# Neuroprotective effect of vitamin B_6_ and vitamin B_12_ against vincristine- induced peripheral neuropathy: A randomized, double- blind, placebo controlled, multicenter trial

**DOI:** 10.1101/2021.05.18.21257296

**Authors:** Fatiha Tasmin Jeenia, Ferdaush Ahmed Sojib, Md Sayedur Rahman, Tasneem Ara, Rafiquzzaman Khan, Md Jamal Uddin Tanin

## Abstract

**Background:** Vincristine leads to development of debilitating neuropathy in 40-45% patients with resultant compromised efficacy of chemotherapy, suboptimal treatment and worse prognostic outcome. Vitamin B_6_ and vitamin B_12_ improves non-oncological neuropathies. Therefore, this study investigated vitamin B_6_ and vitamin B_12_ to prevent vincristine- induced peripheral neuropathy (VIPN) by reducing incidence, absolute risk, relative risk, severity as well as delaying the onset.

**Methods:** Patients with ALL undergoing induction phase were randomly assigned into intervention or placebo arm in a double- blind manner. Vitamin B_6_ (25 mg Pyridoxine) two tablets were given three times daily for 5 weeks. Vitamin B_12_ (500 μg/ml Methylcobalamin) was administered intravenously on day 1, 3 and 5 of every week for 5 weeks during induction period. Placebo arm received oral and intravenous placebo for same duration. Patients were evaluated on the outset of every week by FACT/GOG-NTX questionnaire. Severity was assessed per NCI-CTCAE grading scale.

**Results:** 102 patients were enrolled. Among them 81 completed the study, where 42 received vitamin B_6_ and B_12_ and 39 received placebo. There was significant difference in incidence of neuropathy between arms (26.19% intervention arm, 56.41% placebo; P-0.01). Relative risk of neuropathy was significantly (RR-0.46) lower in intervention arm. Besides, absolute risk reduction (ARR) was 30% and relative risk reduction (RRR) was 54%. NNT was 3.33. Significant trend was observed in difference of severity of VIPN between groups (P-0.03). No significant difference observed in between arms for time to onset of neuropathy.

**Conclusion:** Vitamin B_6_ and vitamin B_12_ significantly reduced the incidence, relative risk and severity of VIPN. NNT was encouraging too. Henceforth, the status of vitamin B_6_ and vitamin B_12_ as neuroprotective agent against VIPN can be recommended as a promising one. Clinicaltrials.gov ID: NCT02923388

## Introduction

Chemotherapy Induced Peripheral Neuropathy (CIPN) is one of the most challenging and complex complications of chemotherapy [1]. CIPN is observed in up to 80% of patients [2]. Chemotherapeutic regimen completion with proper dose and adequate duration is crucial in determining the survival of patients with cancer. Unfortunately, CIPN significantly limits the dose and numbers of chemotherapy and may lead to premature termination of treatment [3]. However, these dosage adjustments affect the prognostic outcome and increase chance of relapse. Moreover, neurotoxicity has negative impact on patient’s quality of life, particularly on daily living activities [4].

Acute lymphoblastic leukemia (ALL) is most common and accounts for three fourths of leukemia cases [5]. Vincristine is an integral constituent of the regime recommended for treatment of acute lymphoblastic leukemia. It is one of the most neurotoxic antineoplastic medicines [6]. Vincristine Induced Peripheral Neuropathy (VIPN) accounts for around 45% of neuropathy incidence [7].

Two clinical trials attempted to explore the efficacy of ACTH (4- 9) analogue in preventing VIPN. The pilot study showed promising result, but a large double blind trial annulled the possibility with its negative result [8,9]. One isolated study evaluated glutamic acid but no recommendations can be drawn [10]. Another trial by Kautio et al., (2009) demonstrated no efficacy of amitriptyline in amelioration of VIPN [11].

Vitamin B_12_ plays a fundamental role in neurological function [12]. It maintains the integrity of neuronal membrane [13,14]. Methylcobalamin is one of the two coenzyme forms of vitamin B_12_. Intravenous methylcobalamin has no interference with the action of other drugs and is relatively well tolerated [15]. However, chemotherapeutic agents may render vitamin B_12_ inert that leads to deficiency [16]. The neurologic sequel is peripheral neuropathy [12] which may lead to decline in physical functions and disability in patients [17]. On the other hand, vitamin B_6_ is an essential cofactor in synthesis of myelin sheath [18]. Besides, vitamin B_6_ has potential antioxidant property [19].

Broadly, VIPN develops as a result of two proposed mechanisms, which include neurodegeneration and oxidative stress [2]. Eventually, both vitamin B_6_ and vitamin B_12_ have significant role in synthetic pathway of myelin sheath and maintenance of its integrity [13,18]. Moreover, these vitamins also got antioxidant activity to some extent [19,20]. Five distinct case reports has been published between 2004 to 2016 figured out the potential efficacy of pyridoxine and pyridostigmine in treating VIPN with cranial involvement [21-25].

Clinical experience shows that CIPN is a difficult problem to deal with. Different strategies to prevent CIPN have been assessed in cell and animal models, though none has reached clinical application. To date no effective strategy to prevent or treat CIPN is available [26]. In the premise of scarce number of RCTs to alleviate VIPN and encouraging results of few case reports generated enthusiasm to conduct the present study. Considering the potentials of vitamin B_6_ and vitamin B_12_, this randomized, double-blind, placebo-controlled multicenter trial was undertaken to investigate the effect of these vitamins in reducing VIPN in ALL patients.

## Materials and Methods

### Study sites and participants

This study was conducted in Bangabandhu Sheikh Mujib Medical University (BSMMU) and Dhaka Medical College Hospital (DMCH). Prior to the study conduct, the research protocol sought approval from the Institutional Review Board (IRB) of Bangabandhu Sheikh Mujib Medical University (BSMMU). The study protocol, questionnaire and informed consent form as well as the methodology of the study was reviewed thoroughly by the institutional review board of BSMMU. Eligibility of each patient was assessed and they were informed about the intervention and the study objectives. They were informed that there is least possible chance of any harm to the patient by inclusion in this study and if any complication arises due to the intervention, they will avail treatment for that complication free of cost. Patient was also informed that they can participate in this study by their free will and also they are free to refuse to participate or to withdraw at any time without compromising their medical care. Patients who took part in the study willingly with written consent interviewed and patients’ confidentiality was strictly maintained. Patient’s personal data regarding name, age, sex and other information was not disclosed anywhere and used for research purpose only.

The study protocol was approved by Institutional Review Board of Bangabandhu Sheikh Mujib Medical University (BSMMU) in compliance with the provision of Declaration of Helsinki and Good Clinical Practice guidelines. This trial was registered in clinicaltrials.gov and trial ID is NCT02923388. Patients were informed about the investigational nature of the study and signed informed written consent.

Newly diagnosed acute lymphoblastic leukemia patients of 18 years or older going to receive induction phase of chemotherapy were eligible for this study. Patients were required to have an ECOG performance status of 0-3. Patients were not allowed on trial if they had a pre-existing peripheral neuropathy of any grade; were receiving anticonvulsants, antidepressants, opioids, vitamin E or any other agents specifically being given to prevent or treat neuropathy. Pregnant women, nursing mothers and patients with head neck tumor were excluded.

### Study design

This study was a randomized, double-blind, placebo-controlled, multicenter trial comparing vitamin B_6_ (pyridoxine) and vitamin B_12_ (methylcobalamin) versus matching placebo for 5 weeks. Randomization, blinding, sequence generation was done prior to patient enrollment.

### Randomization and Sequence Generation

After determining the sample size, patients were randomly allocated into two arms prior to the patient enrollment for the study. Randomization was done by online graph pad software by using computer. Necessary inputs were given to the software regarding sample size, number of sets required and the software automatically generated two distinct sets of random numbers.

Online graph pad calculator also generated the sequence of patient numbers while procreating random numbers and thus equally distributed the patients into two comparable groups. The randomization and sequence generation process was conducted by a competent third person, a distinguished Professor of University who has no relationship with this research. The probability of an individual to receive intervention was made independent of the plausibility of any other individual receiving the same.

### Blinding

Immediately after randomization and sequence generation, random numbers of the two sets were assigned as patient code number. One set was designated as intervention group and another set was placebo group. Then the set of code numbers that belongs to the intervention group were written as patient ID numbers on the packages contained methylcobalamin and pyridoxine. On the other hand, the set belongs to the placebo group were designated as patient ID numbers on the packages contained placebo. This total procedure was conducted by the persons unrelated to this research. Thus the participants, caregiver, outcome assessor and the analyst, who require being blind for such study, were effectively blinded from the knowledge about intervention allocation.

### Allocation Concealment

In order to prevent selection bias, concealment of allocation was done. Third person allocated two distinct sets of random numbers into intervention group and placebo group. After entering into this trial, according to that the participants were assigned into their respective group. This allocated code was recorded in two paper documents and in two separate pen drives, which were sealed within two different envelops. The sealed envelopes containing paper documents and pen drives were preserved to another two distinguished Professors not involved in the randomization and blinding procedure. Thereby, intervention allocation was not known to any person involved in the research in advance or during enrollment.

### Patient Enrollment and Intervention

After taking informed written consent patient enrollment started. Patients on intervention arm were provided with pyridoxin 2 tablets 3 times daily and one ampoule methylcobalamin injection on day1, 3 and 5 of every week for 5 weeks starting from the 1^st^ day of chemotherapy. Each tablet contained 25 mg of pyridoxine and each ampoule contained 500 μg of 1 ml methylcobalamin. On the control arm patients got similar amount of placebo tablet and injection at same duration. Each placebo tablet contained placebo and injection placebo was in similar amber colored ampoule contained 1 ml of normal saline.

### Vincristine Chemotherapy

Patients received vincristine according to standard *Berlin-Frankfurt-Münster* (BFM) protocol and MRC UKALL X protocol at a dose of 1.4 mg/ m^2^ of body weight. Maximum 2 mg of vincristine was administered to a patient according to body weight.

### Patient assessment

Patients were assessed for neuropathy at baseline and at the outset of every week during the induction period for 5 weeks. Development of neuropathy was assessed by 11-item Functional Assessment of Cancer Therapy Neurotoxicity (FACT-NTX) questionnaire. Severity of neuropathy was graded according to National Cancer Institute-Common Toxicity Criteria for Adverse Events (NCI-CTCAE) Neurotoxicity grading scale version 4.

### Outcome measures

The primary outcome measure for this study was to compare incidence of neuropathy between pyridoxine, methylcobalamin treated group and placebo group. Secondary outcome measures were determination of absolute risk, relative risk, risk reduction of neuropathy in pyridoxine, methylcobalamin treated group in comparison to placebo group and comparison of severity and time of onset of neuropathy in between groups.

### Statistical analysis

This study, planned to accrue 102 patients, was designed with 80% power (β= 0.20) to demonstrate a statistical reduction of 25% of the neuropathy events in vitamin B_6_ and vitaminB_12_ treated groups. All the raw data were recorded, processed and analyzed by using computer software SPSS version 23.0. The statistics used to analyze the data were chi-square (x^2^) test, independent t-test, Fisher’s exact test, Kaplan-Meier analysis and log rank test. The chi-square (x^2^) test was employed to compare some patient baseline characteristics and incidence of neuropathy. Data implies independent t-test for analysis were patient age, ECOG performance status, FACT-GOG-NTX neurotoxicity scoring. Furthermore, Fisher’s exact test was performed to compare severity of neuropathy. Kaplan-Meier analysis and log rank test was done to detect the delay in onset of neuropathy. P value <0.05 was considered significant.

## Results

### Accrual and Evaluability

102 patients were accrued. 14 patients were excluded based on the eligibility criteria for the study. Among remaining 88 patients, 47 received intervention and 41 received placebo. 7 patients were dropped out during the research due to the reasons like, DORB (n= 3), denial to take medication (n= 2), death from the disease complication (n=2). For these patients, determination of the study end point was not possible and therefore they were excluded from the per-protocol-analysis. At the end of the research the evaluable patients for neuropathy were a total of 81 [Figure 1].

**Figure 1:**
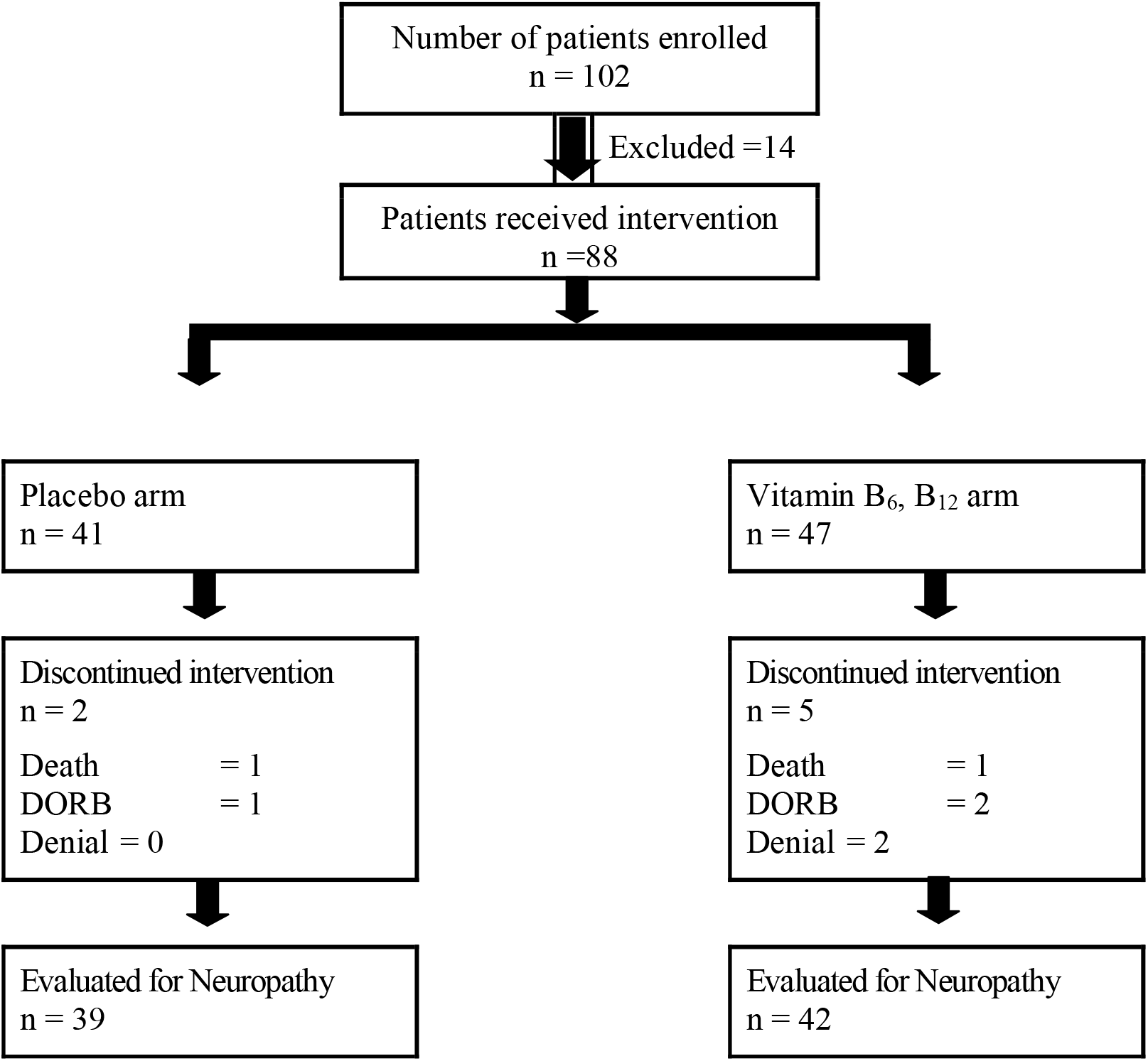
CONSORT diagram.

### Patients Characteristics at Baseline

Table I shows the demographic and clinical characteristics of patients at baseline. There was no significant difference between arms in age, gender and ECOG performance status.

**Table I.**
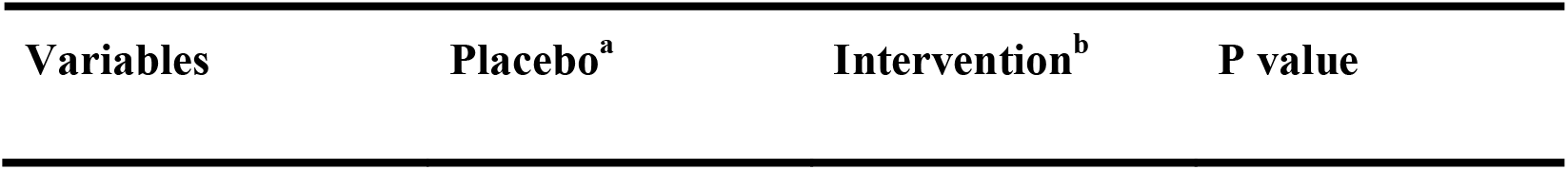
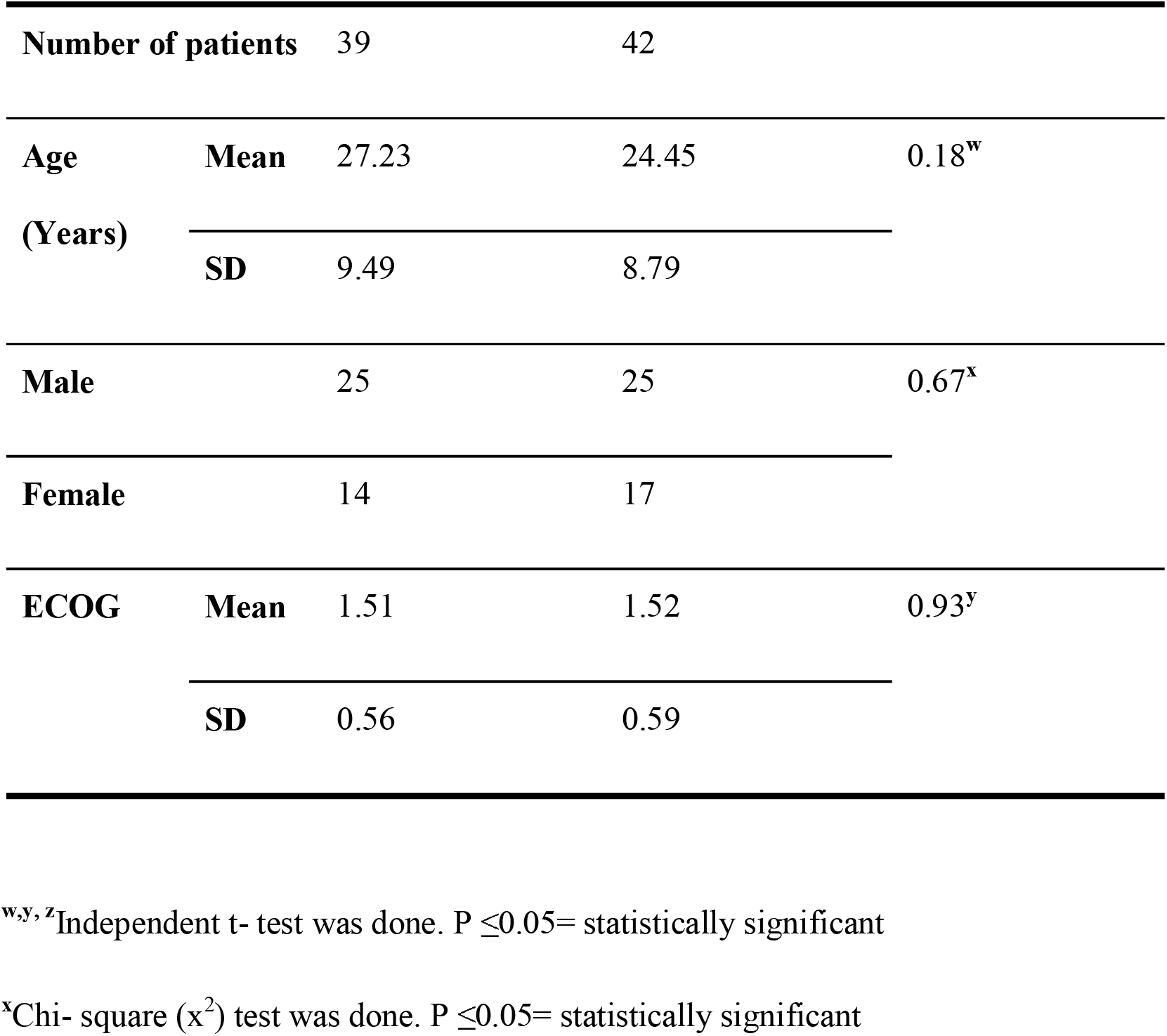
Demographic Characteristics of the Enrolled Patients at Baseline.

| Variables | Placebo<sup>a</sup> | Intervention<sup>b</sup> | P value |
| --- | --- | --- | --- |

|  |  |  |  |  |
| --- | --- | --- | --- | --- |
| **Number of patients** |  | 39 | 42 |  |
| **Age**<br>**(Years)** | **Mean** | 27.23 | 24.45 | 0.18<sup>w</sup> |
|  | **SD** | 9.49 | 8.79 |  |
| **Male** |  | 25 | 25 | 0.67<sup>x</sup> |
| **Female** |  | 14 | 17 |  |
| **ECOG** | **Mean** | 1.51 | 1.52 | 0.93<sup>y</sup> |
|  | **SD** | 0.56 | 0.59 |  |

<sup>w,y,z</sup>Independent t- test was done. $P \leq 0.05$ = statistically significant
<sup>x</sup>Chi- square ( $\chi^2$ ) test was done. $P \leq 0.05$ = statistically significant

### Incidence of Neuropathy

Figure 2 demonstrates that in placebo arm 22 patients out of 39 developed neuropathy, whereas in intervention arm, 11 patients out of 42 developed neuropathy. The incidence of vincristine induced neuropathy in placebo arm was 56.41%, which was 26.19% in intervention arm. Difference in the incidence of neuropathy between placebo and intervention arm was statistically significant (p=0.01).

**Figure 2.**
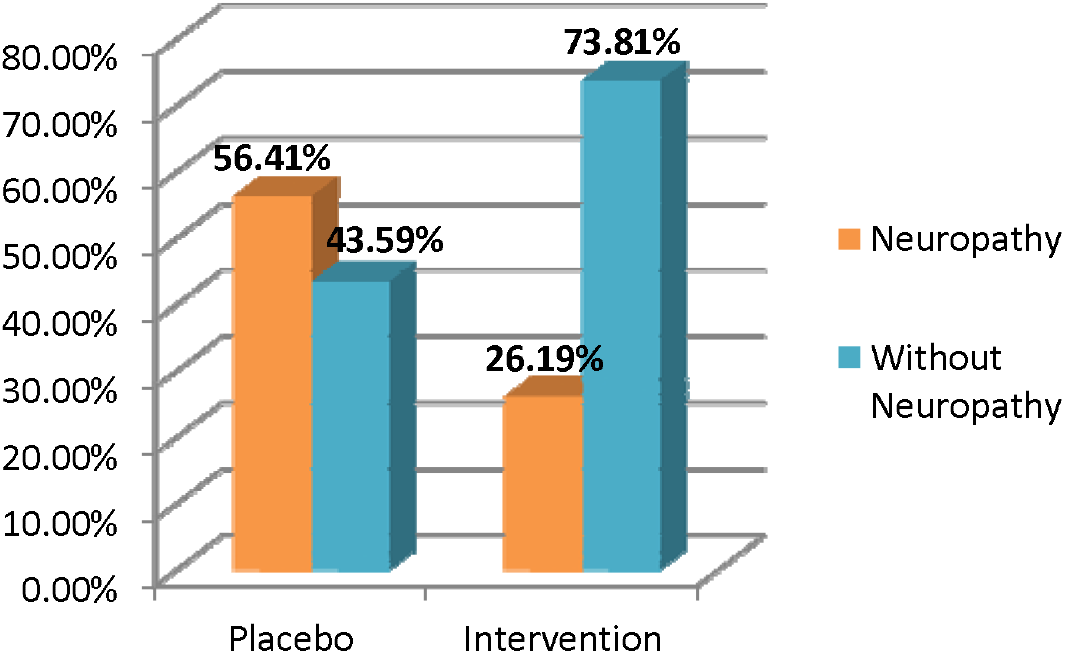
Comparison in Incidence of Neuropathy Between Placebo and Intervention Arms

### Absolute Risk Reduction (ARR), Relative Risk Reduction (RRR) and Number Needed to Treat (NNT) of Placebo and Intervention Arms

Table II demonstrates the Absolute Risk Reduction (ARR),Relative Risk Reduction (RRR) and Number Needed to Treat (NNT) of the given intervention. Absolute Risk Reduction (ARR) for this study is 30%. Relative risk (RR) for this study is found to be less than 1, which indicates that risk of occurrence of neuropathy is significantly less in intervention arm than in placebo arm. Relative risk reduction (RRR) is 54%.The calculated Number Needed to Treat (NNT) is 3.33. Therefore, approximately three patients need to be treated with vitamin B_6_ and vitamin B_12_ in order to prevent neuropathy in one patient.

**Table II.** Absolute Risk Reduction (ARR), Relative Risk Reduction (RRR) and Number Needed to Treat (NNT) of Placebo and Intervention Arms.

|  |  |  |  |
| --- | --- | --- | --- |
| **Absolute risk (AR)** | **Placebo<sup>a</sup> (ARC)** | 0.56 | 56% |
|  | **Intervention<sup>b</sup> (ART)** | 0.26 | 26% |
| **Absolute Risk Reduction (ARR)** |  | 0.30 | 30% |
| **Relative Risk (RR) \*** |  | 0.46 |  |
| **Relative Risk Reduction (RRR)** |  | 0.54 | 54% |
| **Number Needed to Treat (NNT)** |  |  | 3.33 |

\*RR < 1; events are significantly less likely in the intervention group than placebo group

### Neurotoxicity Scoring by FACT/GOG-NTX Subscale Score

The observed values of neuropathy assessment per FACT/GOG-NTX questionnaire and trend of change are shown in Figure 3. On 1^st^ week, the baseline neurotoxicity score difference was not significant between arms (p=0.16). However, on assessment of 2^nd^ week (after 1^st^ dose of vincristine), the score of placebo arm was significantly (p=0.01) lower than intervention arm. Consequently, on assessment of 3^rd^ (p= .01), 4^th^ (p= .03) and 5^th^ week (p= .04), the difference between score of placebo and intervention arm were consistently more significant.

**Figure 3.**
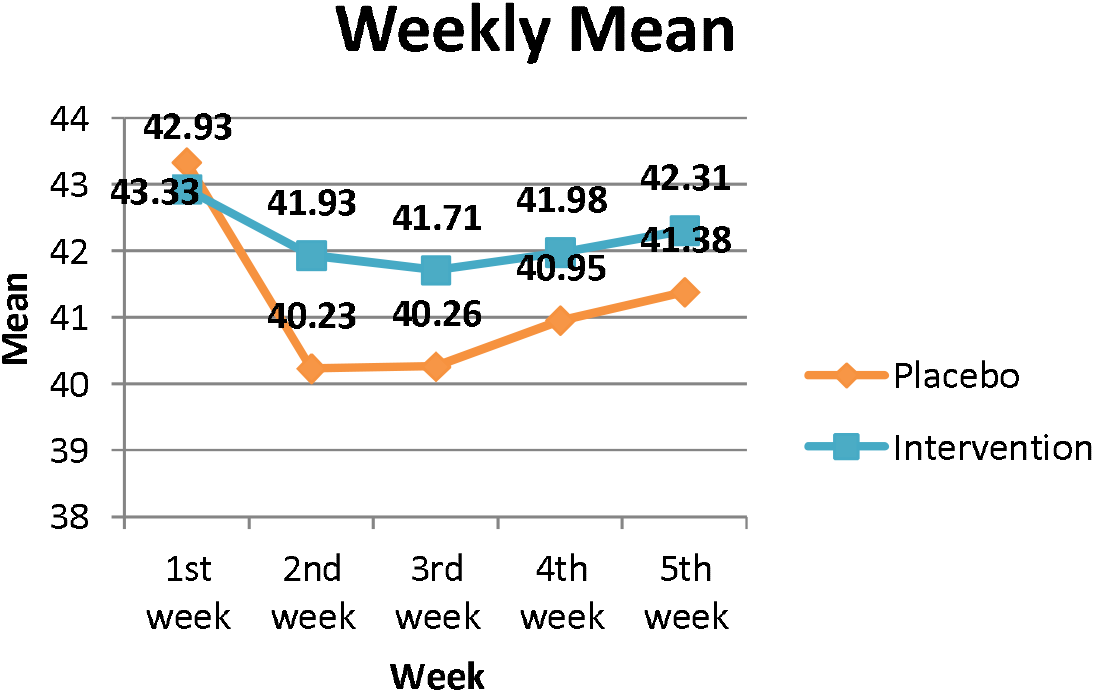
Comparison in the Trend of Change in Neurotoxicity Scoring by FACT/GOG-NTX Subscale Score Between Placebo and Intervention Arms

### Severity of Neuropathy According to NCI-CTCAE Grading Scale

Severity of neuropathy according to NCI-CTCAE scale has been shown in Figure 4. There was statistically significant (p =0.03) difference in the severity of neuropathy between intervention (tablet vitamin B_6_ 50 mg and injection vitamin B_12_ 500 µg) group in comparison to placebo according to NCI-CTCAE grading scale.

**Figure 4.**
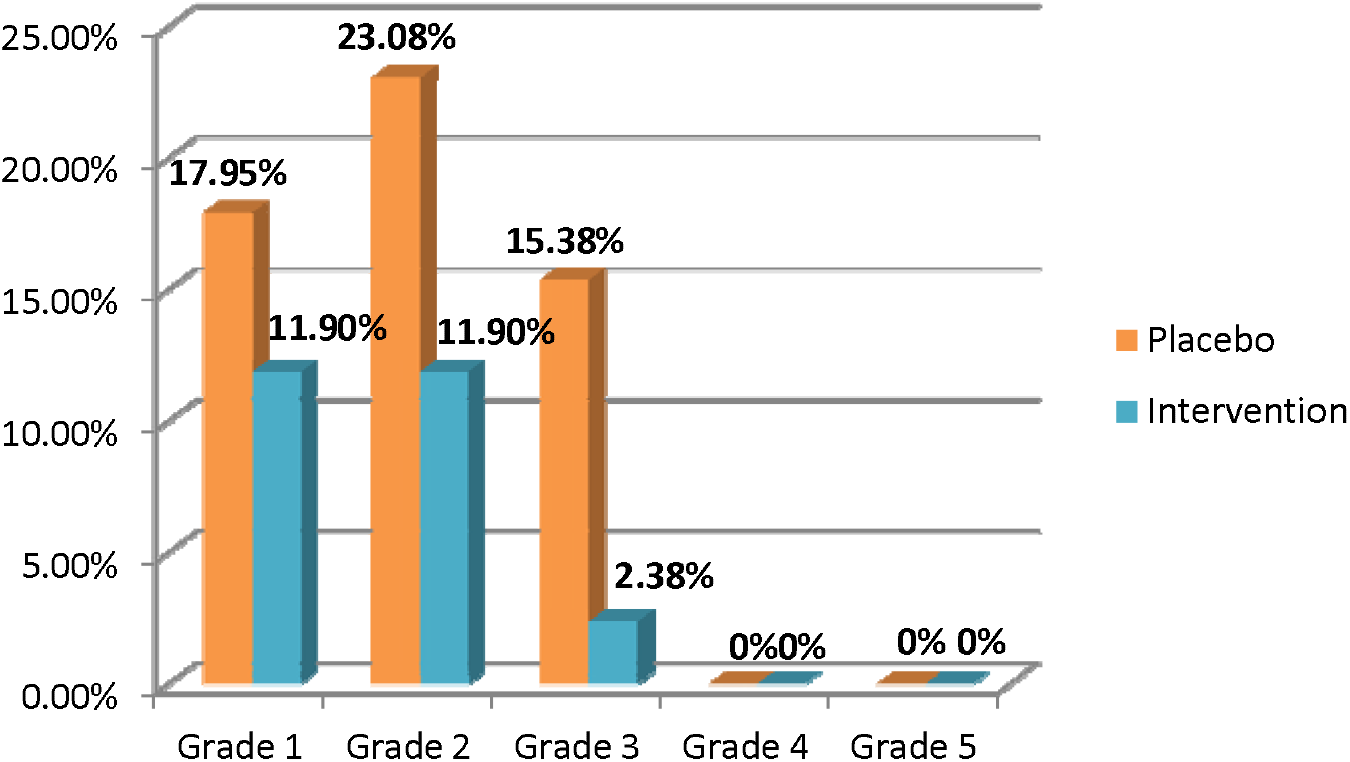
Comparison in Severity of Neuropathy According to NCI-CTCAE Grading Scale Between Placebo and Intervention Arms

### Time of Onset of Neuropathy

The overall comparison in time of onset of Grade 1, 2 and 3 neuropathy between placebo and intervention arms showed no significant difference in the Log Rank (Mantel-Cox) test

## Discussion

Neuroprotection is a tempting alternative to prevent neuropathy induced by cancer chemotherapeutic agents. Review of the existing literatures regarding this alluring option reveals inconsistent results left with almost no recommendation. This trial evaluated the neuroprotective potential of vitamin B_6_ and vitamin B_12_ against vincristine-induced peripheral neuropathy.

One of the primary endpoints of this study was to determine the incidence of neuropathy in intervention and placebo arm. However, the incidence of neuropathy was significantly lower in the patients treated with vitamin B_6_ and vitamin B_12_ (26.19%) in comparison to the patients treated with placebo (56.41%). The incidence of VIPN in placebo arm corresponds with the findings of previous studies like, 43%, 45% and 48% [27,7,28]. The positive findings of previous two studies conducted with vitamin E and omega-3 fatty acid appeared to reduce the incidence of neuropathy in cisplatin and paclitaxel treated patients actually support the finding of the present study [29,30]. Though, the negative finding of two studies conducted with amifostine against incidence of paclitaxel-induced peripheral neuropathy not in inclination with the present study [31,32]. In addition, vitamin E while attempted in patients received taxanes and platinum compounds, there was no significant reduction of neuropathy incidence [33].

Besides the significant result of the present study regarding incidence of neuropathy, the risk reduction analysis which is a powerful tool for interpreting randomized controlled trial also demonstrated significant positive findings. Vitamin B_6_ and vitamin B_12_ resulted into “Absolute Risk Reduction (ARR)” and “Relative risk reduction (RRR)” of vincristine-induced neuropathy by 30% and 54% respectively. The Relative Risk (RR) was 0.46, which indicates significantly lower possibility of developing neuropathy in intervention than placebo. Similar result regarding significant risk reduction by vitamin E and oxcarbazepine was observed in prevention of cisplatin and oxaliplatin-induced peripheral neuropathy [34,35].

Regarding FACT/GOG-NTX questionnaire scoring, the graphical presentation from first week towards second week showed a drastic fall in placebo arm, which is much smaller fall in intervention arm indicating more worsening in placebo arm than intervention. Consequently, significant difference between vitamin B_6_ and vitamin B_12_ treated patients and placebo arm was observed in second, third, fourth and fifth week, which might be explained by the fact that more patients appeared to develop neuropathy with worsening of neuropathic symptoms (Grade 2 and Grade 3) in placebo arm. Guo et al., (2013), who studied with oral alpha lipoic acid to alleviate neuropathy by platinum based compounds, could not reveal any statistically significant difference between placebo and intervention [36]. However, Hershman et al. (2013) reported worsening of CIPN in acetyl-L carnitine treated group indicating contraindication of acetyl -L carnitine in neuropathic symptoms induced by taxanes [37]. The current study revealed safety of vitamin B_6_ and vitamin B_12_ as these vitamins have not worsened vincristine-induced neuropathy.

Another objective of this trial was to compare severity of neuropathy between two arms. Significant difference has been observed in severity of neuropathy between placebo and intervention arms. This particular result is consistent with Milla et al.(2009) who reported significant reduction in severity with glutathione in oxaliplatin treated patients [38]. Aside than that, Ishibashi et al., (2010) where difference of severity of neuropathy between the groups asserted as insignificant when calcium and magnesium was investigated against oxaliplatin-induced neuropathy and findings of Leal et al. (2014) which showed an insignificant reduction of severity by glutathione in paclitaxel and carboliplatin treated patients not in concordance with this present study [39,40].

However, the Kaplan- Meier Curve for the onset of neuropathy corresponds to the log-rank test which showed insignificant difference in the time of onset of neuropathy between two arms. Study finding of Chay et al., (2010) demonstrated an insignificant delay in the time of onset of neuropathy which is compatible with the present study [41]. Grothey et al., (2011) concluded with a significant delay in the onset of neuropathy in intervention arm [42]. These studies were investigating the role of calcium and magnesium in prevention of oxaliplatin-induced neuropathy and were aborted prematurely as calcium and magnesium reported to interfere with the anti tumor efficacy of oxaliplatin. The conceptual strength of this regime is that theoretically as well as in the present study; vitamin B_6_ and vitamin B_12_ did not interfere with the antitumor activity of vincristine.

This study has not evaluated the autonomic neuropathy, a common side effect of vincristine chemotherapy that is more pronounced in adult patients [43]. Risk factors of developing neuropathy like, diabetes mellitus, alcohol consumption, preexisting neuropathy also needs to be taken into consideration [44,45]. According to Quasthoff and Hurtung, (2002), vincristine more than 4mg is considered as the neurotoxic cumulative dose[26]. In this study, all the patients received vincristine at a dose of 1.4 mg/m^2^ with an upper limit of 2 mg and therefore below the recognized neurotoxic dose. Besides, patients with history of diabetes mellitus, neuropathy and alcohol abuse were excluded from the study.

Genetic variation is evident in development of VIPN [46,47]. Thereby, analysis for genetic variant of VIPN might be beneficial to determine the susceptibility of neurotoxicity in this population.

Moreover, this study has not utilized the maximum tolerable dose of vitamin B_6_ and vitamin B_12_. Thereupon, dose escalation might be another preference as both vitamin B_6_ and vitamin B_12_ are water soluble, non-toxic vitamins.

This trial was endeavored with two of the commonly used, safe and inexpensive medicines of peripheral neuropathy with an endurance to generate a potential candidate for preventing VIPN. In this study, vitamin B_6_ and vitamin B_12_ have explicitly and unquestionably reduced the incidence, severity and the risk of vincristine-induced neuropathy in terms of both absolute and relative risk.

## Data Availability

All data are available

## Acknowledgment

Heartfelt gratitude to authority of BSMMU for providing fund for this research. I would like to express my gratitude to Prof. Ahmed Abu Saleh, Department of Microbiology of BSMMU and Prof. Humayun Sattar, Chairman of the Department of Microbiology of BSMMU for their gracious help regarding randomization and blinding procedures of this research.

## References

1. Boland BA, Sherry V, Polomano RC. Chemotherapy-Induced Peripheral Neuropathy in Cancer Survivors. Oncology (Nursing Edition) 2010; 24: e1-3. Available at: http://www.cancernetwork.com/oncology-nursing/chemotherapy-induced-peripheral-neuropathy-cancer-survivors [Accessed on 19th February, 2020]

2. Sisignano M, Baron R, Scholich A, Geisslinger G. Mechanism-based treatment for chemotherapy-induced peripheral neuropathic pain. Nat Rev Neurol 2014. 10: 694–707.

3. Wang XM, Lehky TJ. Discovering Cytokines as Targets for Chemotherapy-Induced Painful Peripheral Neuropathy. Cytokine 2012. 59: 3–9.

4. Driessen CML, de Kleine-Bolt KME, Vingerhoets AJJM, Mols F, Vreugdenhil G. Assessing the impact of Chemotherapy-induced peripheral neurotoxicity on the quality of life of cancer patients. Support Care Cancer 2012; 20: 877–81.

5. American Cancer Society. Cancer Facts and Figures 2012. Available from: https://www.cancer.org/content/dam/cancer-org/research/cancer-facts-and-statistics/annual-cancer-facts-and-figures/2012/cancer-facts-and-figures-2012.pdf [Accessed on 19th February, 2020]

6. Argyriou AA, Bruna J, Marmiroli P, Cavaletti G. Chemotherapy-induced peripheral neurotoxicity (CIPN): An update. Critical Reviews in Oncology/Hematology 2012; 82: 51–7.

7. Rea D, Legros L, Raffoux E, Thomas X, Turlure P, Maury X, et al. High-dose imatinib mesylate combined with vincristine and dexamethasone (DIV regimen) as induction therapy in patient with resistant Philadelphia-positive acute lymphoblastic leukemia and lymphoid blast crisis of chronic myeloid leukemia. Leukemia 2006. 20: 400–3.

8. Van Kooten B, Van Diemen HAM, Groenhout KM, Huijgens PC, Ossenkoppele GJ, Nauta JJP, et al. A pilot study on influence of a corticotrophin (4-9) analogue on vinca alkaloid-induced neuropathy. Arch Neurol 1992. 49: 1027–31.

9. Koeppen S, Verstappen CCP, Korte R, Scheulen ME, Strumberg D, Postma TJ, et al. Lack of neuroprotection by an ACTH (4-9) analogue: A randomized trial in patients treated with vincristine for Hodgkin’s or non-Hodgkin’s lymphoma. J Cancer Res Clin Oncol 2004. 130: 153–60.

10. Jackson DV, Wells HB, Atkins JN, Zekan PJ, White DR, Richards F, et al. Amelioration of vincristine neurotoxicity by glutamic acid. Am J Med 1988. 84: 1016–22.

11. Kautio AL, Haanpaa M, Leminen A, Kalso E, Kautiainen H, Saarto T. Amitriptyline in the prevention of chemotherapy-induced neuropathic symptoms. Anticancer Research 2009. 29: 2601–6.

12. Oh RC, Brown DL. Vitamin B_12_ Deficiency. Am Fam Physician 2003. 67: 979–86.

13. Singh NN, Thomas FP, Diamond AL. Vitamin B_12_ Associated Neurological Diseases. Medscape. Available from: http://emedicine.medscape.com/article/1152670-overview#showall [Accessed on 19th February, 2020]

14. Malouf R, Sastre AA. Vitamin B12 for cognition (Review). Cochrane library 2003. 1–12.

15. Sweetman SC. (ed.) Nutritional Agents and Vitamins. In: Martindale The Complete Drug Reference. 36th ed. London, UK: Pharmaceutical Press; 2009. pp. 1981–3.

16. Garcia A, Tisman G. Metformin, B12 and Enhanced Breast Cancer Response to Chemotherapy. J Clin Oncol 2010, 28: e19.

17. Leishear K, Boudreau RM, Studenski SA, Ferrucci L, Rosano C, de Rekeneire N, et al. The Relationship of Vitamin B_12_ and Sensory and Motor Peripheral Nerve Function in Older Adults. J Am Geriatr Soc 2012. 66: 1057–63.

18. Frye RE, Jabbour SA. Pyridoxine deficiency. In, Drugs & Diseases (Endocrinology). Medscape. Available from: http://emedicine.medscape.com/article/124947-overview#showall [Accessed on 19th February, 2020]

19. Higashi-Okai K, Nagino H, Yamada K, Okai Y. Antioxidant and prooxidant activities of B group vitamins in lipid peroxidation. J UOEH 2006. 28: 359–68.

20. Pahlavanzadeh F, Bidadkosh A, Derakhshanfar A, Rastegar AM, Rushanzamir M. Antioxidant protecting effects of vitamin B6 at reducing hemodynamic toxicity of gentamycin in rat model of nephrotoxicity. Comp Clin Pathol, 2013. 22: 637–43.

21. Karaman K, Akbayram S, Garipardic M, Oner AF. Successful treatment of vincristine induced unilateral ptosis with pyridoxine and pyridostigmine in a child with Langerhans cell histiocystosis (LCH). Eur J Gen Med 2016. 13: 67–9.

22. Bhat KG, Singhal V, Borker AS. Successful treatment of vincristine induced ptosis and polyneuropathy with pyridoxine and pyridostigmine in a child with acute lymphoblastic leukemia. Indian J Med Pediatr Oncol 2012; 33: 185–7.

23. Ozyurek H, Turker H, Akbalik M, Bayrak AO, Ince H, Duru F. Pyridoxine and pyridostigmine treatment in vincristine-induced neuropathy. Pediatr Hematol Oncol 2007. 24: 447–52.

24. Bay A, Yilmaz C, Yilmaz N, Oner AF. Vincristine induced cranial polyneuropathy. Indian Pediatr 2006; 73: 531–3.

25. Muller L, Kramm CM, Tenenbaum T, Wessalowski R, Gobel U. Pediatr Blood Cancer 2004. 42: 287–8.

26. Quasthoff S, Hartung HP. Chemotherapy-induced peripheral neuropathy. J Neurol, 2002. 249: 9–17.

27. Powles TJ, Jones AL, Judson IR, Hardy JR, Ashley SE. A randomized trial comparing combination chemotherapy using mitomycin C, mitozantone and methotrexate (3M) with vincristine, anthracycline, and cyclophosphamide (VAC) in advanced breast cancer. Br J Cancer 1991. 64: 406–10.

28. Jackson DV, Paschold EH, Spurr CL, Muss HB, Richards F, Cooper MR. Treatment of advanced non-Hodgkin’s lymphoma with vincristine infusion. Cancer 1984. 53: 2601–6.

29. Pace A, Savarese A, Picardo M, Maresca V, Pacetti U, Monte GD, et al.Neuroprotective effect of vitamin E supplementation in patients treated with cisplatin chemotherapy. J Clin Oncol 2003. 21: 927–31.

30. Ghoreishi Z, Esfahani A, Djazayeri A, Djalali M, Golestan B, Ayromlou H, et al. Omega-3 fatty acids are protective against paclitaxel-induced peripheral neuropathy: A randomized, double-blind, placebo controlled trial. BMC Cancer 2012. 12: 355.

31. Leong SS, Tan EH, Fong KW, Wilder-Smith E, Ong YK, Tai BC, et al. Randomized, double-blind, trial of combined modality treatment with or without amifostine in unresectable stage III non small cell lung cancer. J ClinOncol 2003. 21: 1767–74.

32. Openshaw H, Beamon K, Synold TW, Longmate J, Slatkin NE, Doroshow JH, et al. Neurophysiological study of neuropathy after high-dose paclitaxel: Lack of neuroprotective effect of amifostine. Clin Cancer Res 2004. 10: 461–7.

33. Kottschade LA, Sloan JA, Mazurczak MA, Johnson DB, Murphy BP, Rowland KM, et al. The use of vitamin E for prevention of chemotherapy-induced peripheral neuropathy: Results of a randomized, phase III clinical trial. Support Care Cancer 2011. 19: 1769–77.

34. Argyriou AA, Chroni E, Koutras A, Iconomou G, Papapetropoulos S, Polychronopoulos P, et al. A randomized controlled trial evaluating the efficacy and safety of vitamin E supplementation for protection against cisplatin-induced peripheral neuropathy: Final results. Support Care Cancer 2006; 14: 1134–40.

35. Argyriou AA, Chroni E, Polychronopoulos P, Iconomou G, Koutras A, Makatsoris T, et al. Efficacy of oxcarbazepine for prophylaxis against cumulative oxaliplatin-induced neuropathy. Neurology 2006; 67: 2253–5.

36. Guo Y, Jones D, Palmer JL, Forman A, Dakhil SR, Velasco MR, et al. Oral alpha-lipoic acid to prevent chemotherapy-induced peripheral neuropathy: A randomized, double-blind, placebo-controlled trial. Support Care Cancer 2013. 22: 1223–31.

37. Hershman DL, Unger JM, Crew KD, Minasian LM, Awad D,Moinpour CM, et al. Randomized double-blind placebo-controlled Trial of Acetyl-L-Carnitine for the prevention of taxane-induced neuropathy in women undergoing adjuvant breast cancer therapy. J Clin Oncol 2013. 31: 2627–33.

38. Milla P, Airoldi M, Weber G, Drescher A, Jaehde U, Cattel L. Administration of reduced glutathione in FOLFOX4 adjuvant treatment for colorectal cancer: effect on oxaliplatin pharmacokinetics, pt-DNA adduct formation, and neurotoxicity. Anti-Cancer Drugs, 2009. 20: 396–402.

39. Ishibashi K, Okada N, Miyazaki T, Sano M, Ishida H. Effect of calcium and magnesium on neurotoxicity and blood platinum concentrations in patients receiving mFOLFOX6 therapy: a prospective randomized study. Int J Clin Oncol 2010. 15, pp. 82–7.

40. Leal AD, Qin R, Atherton PJ, Haluska Jr P, Behrens RJ, Tiber C, et al. NCCTG N08CA (Alliance): The use of glutathione for prevention of paclitaxel/carboliplatin induced peripheral neuropathy: A phase III randomized, double-blind, placebo controlled study. Cancer 2014. 120: 1890–97.

41. Chay WY, Tan SH, Lo YL, Ong SYK, Ng HC, Gao F, et al. Use of calcium and magnesium infusions in prevention of oxaliplatin induced sensory neuropathy. Asia-Pac J Clin Oncol 2010; 6: 270–7.

42. Grothey A, Nikcecich DA, Sloan JA, Kugler JW, Silberstein PT, Dentchev T, et al. Intravenous calcium and magnesium for oxaliplatin-induced sensory neurotoxicity in adjuvant colon cancer: NCCTG N04C7. J Clin Oncol 2011. 29: 421–7.

43. Imam EA, Ibrahim A, Palaian S, Ibrahim MIM. Prevalence of vincristine-induced peripheral neuropathy among Sudanese cancer patients. J Young Pharm 2016. 8: 239–43.

44. Kawakami K, Tunoda T, Takiguchi T, Shibata K, Ohtani T, Kizu J, et al. Factors exacerbating peripheral neuropathy induced by paclitaxel plus carboliplatin in non-small cell lung cancer. Oncology Research 2012. 20: 179–85.

45. Dimopoulos MA, Mateos MV, Richardson PG, Schlag R, Khuageva NK, Shpilberg O, et al. Risk factors of, and reversibility of, peripheral neuropathy associated with bortezomib-melphalan-prednisolone in newly diagnosed patients with multiple myeloma: sub-analysis of the phase 3 VISTA study. European J Haem 2011; 86: 23–31.

46. Egbelakin A, Ferguson MJ, MacGill EA, Lehmann AS, Topletz AR, Quinney SK, et al. Increased risk of vincristine neurotoxicity associated with low CYP3A5 expression genotype in children with acute lymphoblastic leukemia. Pediatr Blood Cancer 2011, 56: 361–7.

47. Dennison JB, Kulanthaivel P, Barbuch R, Renbarger JL, Ehlhardt WJ, Hall SD. Selective metabolism of vincristine in vitro by CYP3A5. DMD 2006; 34: 1317–27.

